# Estimating the time-varying reproduction number of COVID-19 with a state-space method

**DOI:** 10.1101/2020.07.09.20150219

**Authors:** Shinsuke Koyama, Taiki Horie, Shigeru Shinomoto

## Abstract

After slowing down the spread of the novel coronavirus COVID-19, many countries have started to relax their confinement measures in the face of critical damage to socioeconomic structures. At this stage, it is desirable to monitor the degree to which political measures or social affairs have exerted influence on the spread of disease. Though it is difficult to trace back individual transmission of infections whose incubation periods are long and highly variable, estimating the average spreading rate is possible if a proper mathematical model can be devised to analyze daily event-occurrences. To render an accurate assessment, we have devised a state-space method for fitting the Hawkes process to a given dataset of daily confirmed cases. The proposed method detects changes occurring in each country and assesses the impact of social events in terms of the temporally varying reproduction number, which corresponds to the average number of cases directly caused by a single infected case. Moreover, the proposed method can be used to predict the possible consequences of alternative political measures. This information can serve as a reference for behavioral guidelines that should be adopted according to the varying risk of infection.

## INTRODUCTION

While the novel coronavirus COVID-19 has spread worldwide, different countries have employed various intervention strategies, many of which were later followed by liberalized approaches and relaxed behaviour from individuals. In this situation, it is desirable to monitor the extent to which individual political measures have influenced the spread of the disease in each country and predict the possible consequences of alternative measures.

A fundamental metric representing the degree of the spread of a disease is the reproduction number, *R*, which is defined as the average number of cases directly caused by a single case [1, 2]. However, it is difficult to trace the concrete processes by which infections have been transmitted among individual people, particularly considering the protection of private information. Thus, a statistical analytical method is needed to infer the underlying process from the available data consisting of the number of daily infected cases, which were obtained by imperfect observation and accompanied by errors.

Mathematical epidemiological studies using the ordinary differential equation (ODE) models, such as the susceptible-infectious-recovered (SIR) model, have contributed to our understanding of causal factor dynamics, the results of which can be used to suggest control measures needed in given situations [3-5]. While the original study of Kermack and McKendrick in 1927 [6] considered the distribution of delays in the transmission of a disease, the majority of later studies used ODEs in favor of an analytical treatment [7]. Though ODE models also assume the transmission delay, they cannot consider the distribution of delays, which are widely dispersed from 2 to 14 days in the case of COVID-19 [8–11]. To capture the time-varying reproduction number under fluctuating circumstances, it is necessary to incorporate the delay distribution explicitly in the analysis, as previously performed in an analysis using the semi-mechanistic Bayesian hierarchical model [12].

Herein, we establish a state-space method for estimating the time-varying reproduction number by fitting the Hawkes process [13], which explicitly adopts the delay distribution. While the semi-mechanistic Bayesian hierarchical model [12] requires manual assignment of the change-points, our method automatically detects the change-points solely from a given series of the number of daily cases. We first apply the method to synthetic data to confirm that the method properly detects the changepoints embedded in the simulation. Here, the proposed method is compared with a conventional method in terms of performance estimation of the time-varying reproduction ratio. Then, we apply the proposed method to real data and examine whether the detected changes are consistent with the times at which political measures had been implemented in each country. The proposed method can also predict the number of new cases in the future to examine any possible consequences of alternative political measures.

## RESULTS

### Hawkes process

As a basic model describing the transmission of disease, we adopt the Hawkes process [13], which describes a self-excitation process in terms of the instantaneous occurrence rate λ(*t*) as

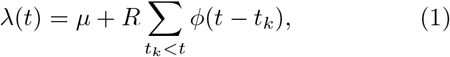

where *μ* is the spontaneous occurrence rate, and the second term represents a self-excitation effect such that the occurrence of an event adds the probability of future events (Fig. 1). *R* is the reproduction number representing the average number of events induced by a single event, *t_k_* is the occurrence time of a past (fcth) event, and *ϕ*(*t*) is a kernel representing the distribution of the transmission delays, satisfying the normalization 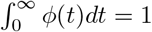. Events {*t*_1_, *t*_2_, ⋯} are derived randomly in time from the rate λ(*t*).

**FIG. 1.**
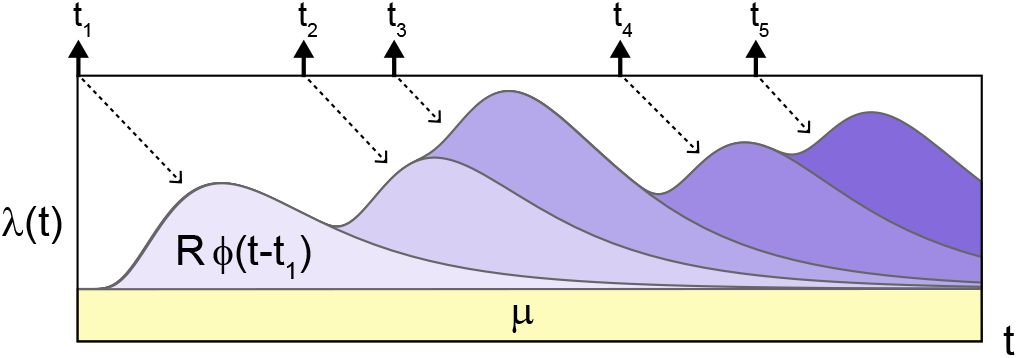
Schematic description of the Hawkes process Eq. (1). The occurrence rate *λ*(*t*) is increased according to past events occurred at times *t* = *t_k_* (*k* = 1; 2; *⋯*) with the transmission delays *t−t_k_* distributed with *ϕ*(*t − t_k_*). *R* is the reproduction number that represents the average number of events induced by a single event.

We convert the original instantaneous rate, Eq. (1), into the expected number of events on a daily basis:

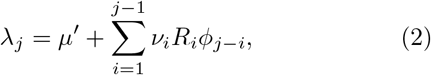

where *λ_j_* is the expected number of events on *j*th day. The first term *μ*′ on the right-hand-side refers to the expected number of spontaneous occurrences on a daily basis. The second term represents the self-excitation process, the manner in which *v_i_* events that have occurred on a day *i* exerted influence with the delay of *j − i* days. Here, we assume that the reproduction number may change and represent the daily dependence as {*R_i_*}*_i_. ϕ_j−i_* represents a distribution of the transmission delays *d* = *j − i*, satisfying the normalization 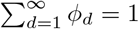. The number of events on each day *v_i_* is derived from the distribution characterized by the expected number on that day *λ_i_* (METHODS).

The COVID-19 model parameters were chosen as follows: the spontaneous occurrence of infection is absent, *μ*′ = 0, because there is no spontaneous occurrence for COVID-19 except at the initial occurrence in China. The virus is transmitted between individuals during close contact, and each individual is determined to have an episode of infection. The duration between symptom onsets of successive cases is referred to as the serial interval [14], which is slightly different from the incubation period [15, 16]. It is reported that the distribution of the serial intervals is suitably approximated with the lognormal distribution function of the mean 4.7 days and SD 2.9 days for COVID-19 [11]. We have adopted this distribution as the transmission delay kernel *ϕ_d_*.

Given a series of the time-varying reproduction number {*R_i_*}*_i_*, we can obtain the number of events {*v_i_*}*_i_* with Eq. (2). Because the generation process contains randomness in deriving the number of events *v_i_* from *λ_i_*, however, we cannot trace back the process straightforwardly. To estimate the time-varying reproduction number {*R_i_*}*_i_* from the number of daily confirmed cases {*v_i_*}*_i_*, we have constructed a Bayesian state-space method that fits the converted Hawkes process to a given dataset (METHODS).

### Analysis of synthetic data

Firstly, we evaluated the functionality of the state-space method by applying it to synthetic data. For this purpose, we constructed simulations of the Hawkes process mimicking prototypical evolutions in several countries. In the simulations, we took *μ’* = 0 and began with a few infections as initial seeds, mimicking those who introduced the disease into each country. With an initial reproduction number *R >* 1, the daily cases initially grew exponentially. To reproduce a variety of evolutions in different countries, we have evaluated several schedules of the reproduction number {*R_i_*}*_i_*.

Figure 2 depicts three prototypical cases: the rapid increase is followed by a slow decrease, as in Italy and France (type A); the rapid increase is followed by a rapid decrease, and then it started to increase again, as in Japan, and Australia (type B); and the increase is followed by a decrease, and then another large increase, as in Iran and Saudi Arabia (type C). For each type of time-varying reproduction number {*R_i_*}*_i_*, the Hawkes process was simulated over an interval of length *T* = 120 days to generate daily cases {*v*_1_, *…, v_T_*} (Fig. 2).

**FIG. 2.**
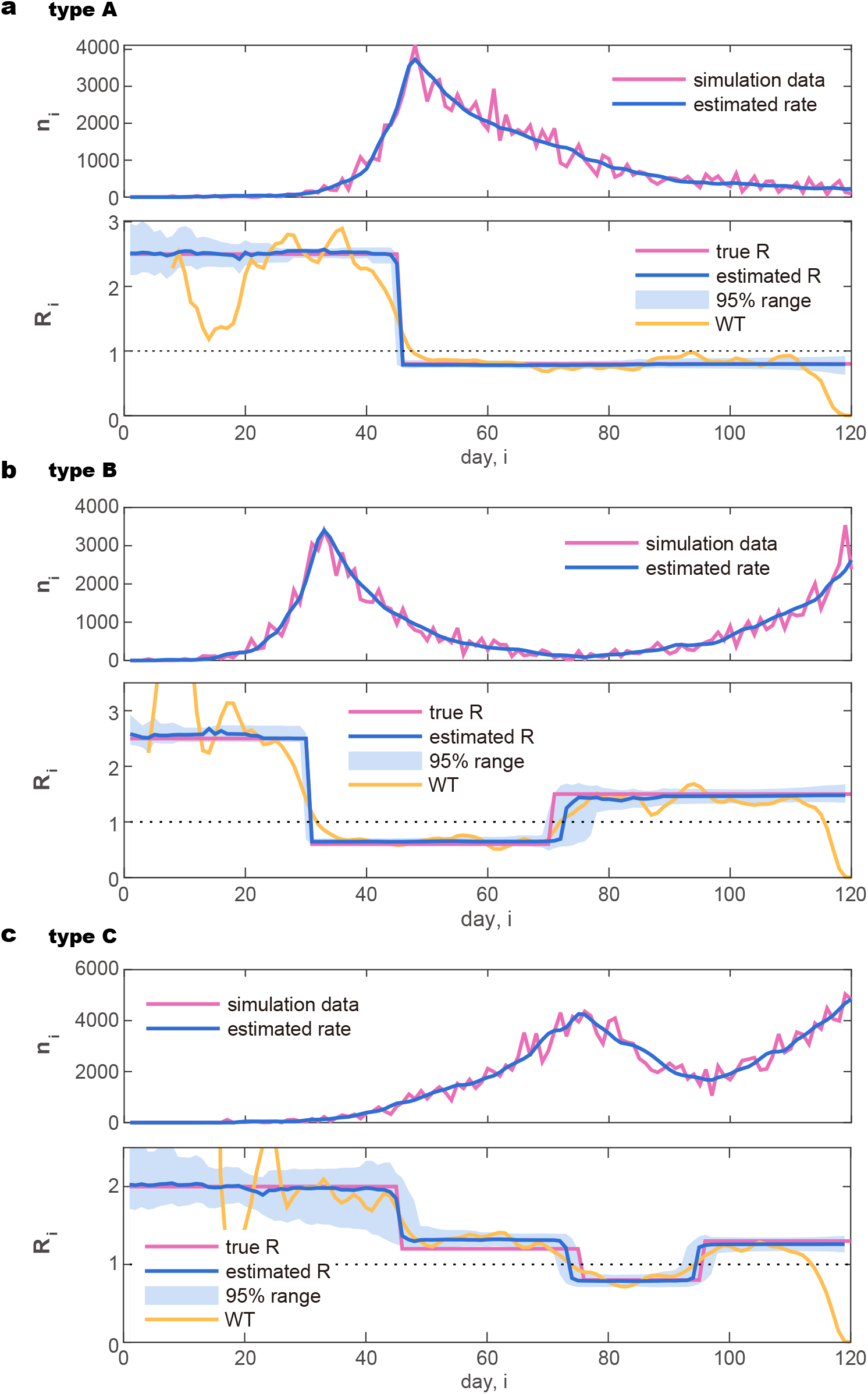
Synthetic daily cases obtained by simulating the Hawkes process and the estimated reproduction number. (a) Type A: rapid increase followed by a slow decrease. (b) Type B: increase followed by a rapid decrease, and then an increase. (c) Type C: slow increase followed by a decrease, and then another large increase. In the upper panel plotting the number of daily cases (purple line), the rate estimated by the state-space method 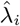 is also plotted (blue line). In the lower panel, the reproduction number *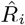* estimated with the state-space method is plotted in reference to the true reproduction number *R_i_* (purple line). The blue solid line and the shaded area represent the median and 95 % range of the posterior distribution, respectively. The reproduction number estimated by a conventional method suggested by Wallinga and Teunis is also plotted for reference (WT: orange line).

We have applied the state-space method to each series of daily new cases; we performed the sequential Monte Carlo algorithm with 10^6^ particles to compute the posterior distributions of the reproduction number for each day, 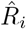. Figure 2 depicts the median (solid line) and 95% range (shaded areas) of the posterior distributions. We see that the amplitude of the reproduction number is estimated properly. In particular, the method has successfully detected change-points in {*R_i_*}*_i_* that were embedded in the simulation.

We compared our method with a conventional estimation method by applying them to the synthetic data. The method suggested by Wallinga and Teunis (WT method) is currently accepted as a standard tool for estimating the reproduction number [17]. It estimates the relative likelihood that a case is infected by another case, given their difference in time of symptom onset. We have also plotted the estimation results obtained with the WT method in Fig. 2. It is observed that the WT method is easily influenced by the fluctuation of data and accordingly it is difficult to discern the change points in the original process. Furthermore, the WT method tends to underestimate the current reproduction number at the end of the recorded interval, because it requires data that could be obtained in the future.

### Analysis of real data

Next, we apply the state-space method to real data of daily confirmed cases in several countries. The number of daily new cases in various countries is made available on websites hosted by public research centers such as Our World in Data (https://ourworldindata.org/coronavirus-source-data) and the Humanitarian Data Exchange (https://data.humdata.org/dataset/novel-coronavirus-2019-ncov-cases). We used data from the former site in this analysis.

#### Variation by day of the week

In the number of reported infections, a large variation has been observed by day of the week; reported infections tend to be fewer on the weekend than on the weekdays. There might have been variations in the original infectious activity due to human behavior, but it is more likely that this variation was caused by the delay in confirming infections and compiling the results at the weekend. The variation by day of the week is commonly observed, but there are large differences between countries, presumably due to the cultural difference in weekly activities (Fig. 3).

**FIG. 3.**
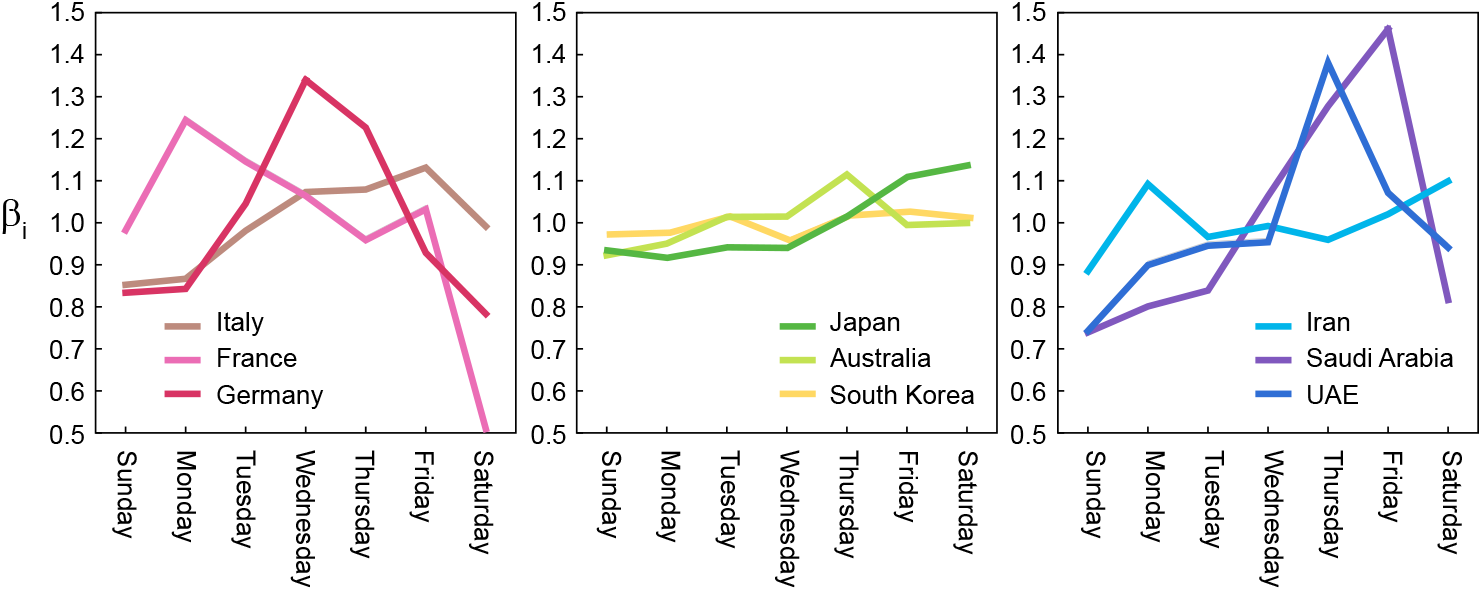
Variations by day of the week in the number of reported infections {*β_i_*} computed for several countries.

Before analyzing a sequence of daily cases of a given country, we process the data as follows; we first obtained the gross daily variation *β_i_* in a week by averaging over the entire infection record (from March 1 2020 to the present), so that the average over a week is normalized as

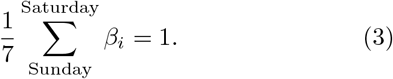

Then, we convert the original data of daily infections {*n_1_,…, n_T_*} to an adjusted dataset {*v*_1_*,…, v_T_*} by

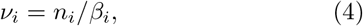

to which we apply the proposed state-space method.

In Fig. 4, we show original daily new infection cases {*n*_1_,…, *n_T_*} and the adjusted dataset {*v*_1_,…, *v_T_*} of several countries. Below each panel of the daily cases, we demonstrate the reproduction number 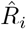 estimated with the proposed state-space method:

**FIG. 4.**
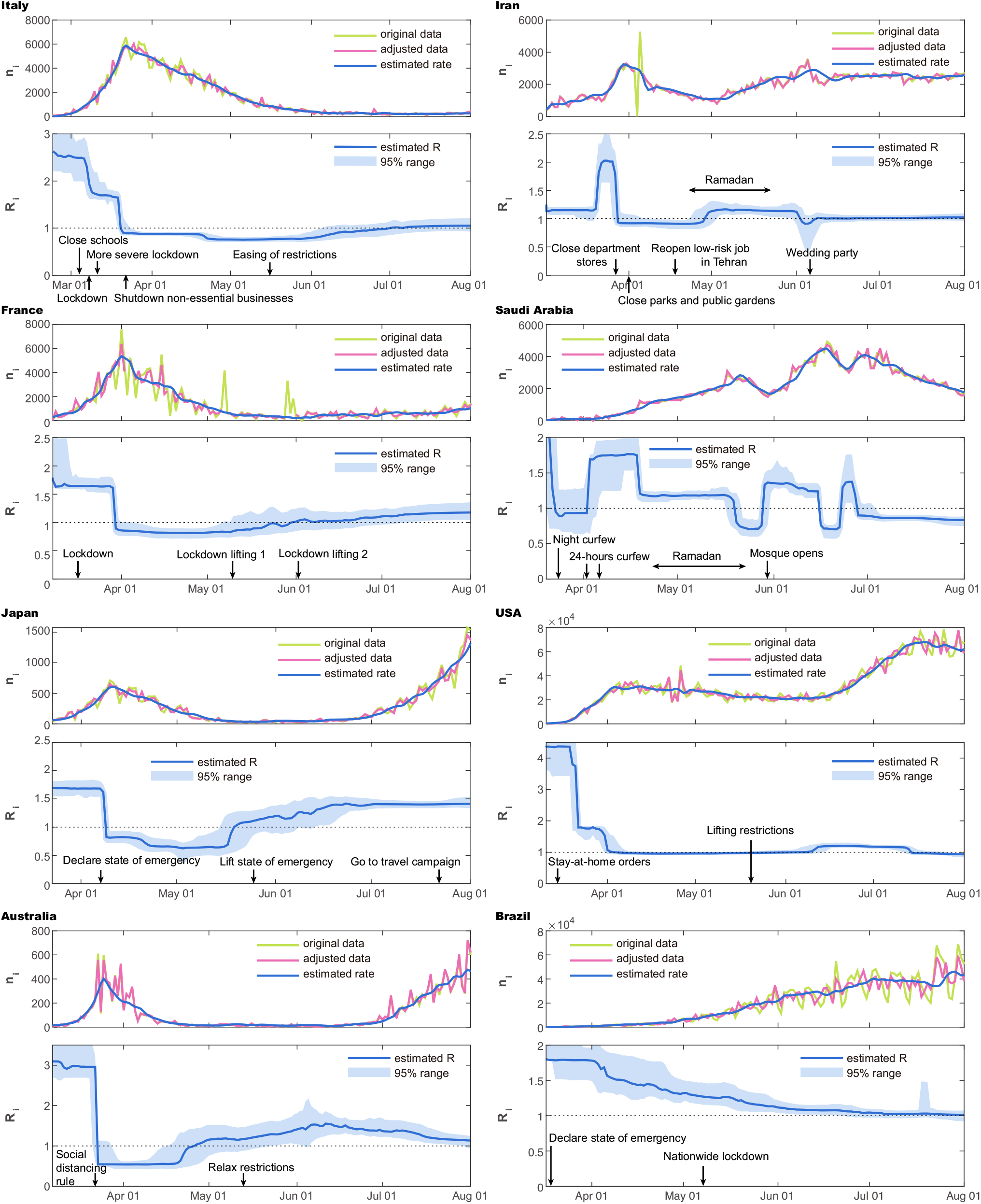
Number of daily new cases and the reproduction number 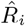 estimated using the state-space method. (left) Italy, France, Japan, and Australia; (right) Iran, Saudi Arabia, and the USA, and Brazil.

> A rapid increase in new cases was followed by a slow decrease. The estimated reproduction number was 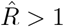 at the outset and dropped to 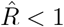. It is interesting to note that the drop in the reproduction number occurred after political measures, such as lockdown and border closure, were enforced.

> The number of cases in countries of Asia and Oceania is found to be relatively small compared to those in Europe. An increase in the number of cases was followed by a rapid decrease, and then by a second increase. Accordingly, the reproduction number exhibited a drop from 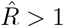 to 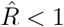, and then it increased to 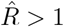.

> The number of new cases in these countries alternately moved up and down, and so accordingly, the estimated reproduction number 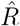 repeatedly crossed the value unity. It can be observed that Ramadan has promoted increased reproduction number, as it may have facilitated human contact.

> A rapid increase of new cases is followed by a very slow decrease, and then another growth. The estimated reproduction number 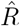 was higher than unity at the beginning, dropped off to near unity due to the confinement measures taken, but then it exceeded unity again. The political measures taken were found to vary by state, making it difficult to interpret the data from this country as a whole.

> The number of new cases keeps growing. The estimated reproduction number 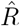 has remained greater than unity.

We also compared the proposed method with a conventional WT method [17] by applying both of them to these real data (Appendix). It is also observed that the WT method is easily influenced by the fluctuation of data.

#### The reproduction numbers at the initial phase

There have been debates about why infection rate and mortality rate change by orders of magnitude across different countries. Though these numbers likely reflect the confinement measures taken in individual countries, there might also have been differences across nations in susceptibility to COVID-19, reflecting not only genetic resistance but also lifestyle and cultural differences, such as shaking hands or hugging.

Because most governments did not implement serious confinement measures at the initial phase, the initial exponential increase of infections might reflect the natural susceptibility of citizens of each country. We realized that the estimated reproduction number was stable in a certain period before each country took confinement measures such as a lockdown or social distancing. Figure 5a depicts the reproduction numbers estimated with the proposed state-space method for 10 days until the day before the confinement measures of each country. The initial variation in the numbers of daily new cases is depicted in Fig. 5b, indicating that the estimated reproduction number is correlated to the slope in the log plot. Here we have selected the period shifted by 5 days, by taking account of the typical transmission delays. We can observe that countries in different regions tend to cluster, indicating that the susceptibility tended to be similar between nations in the same region.

**FIG. 5.**
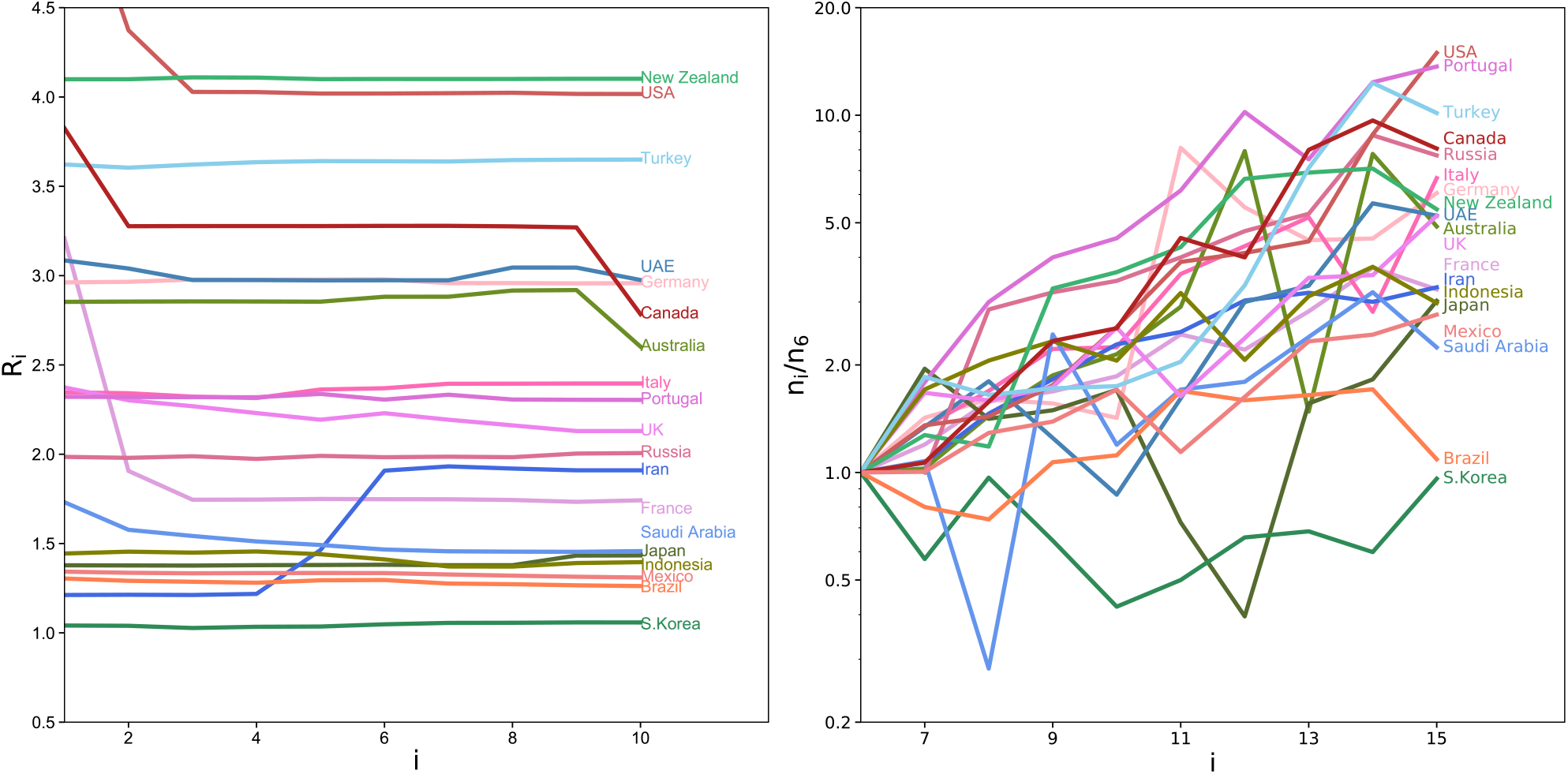
The difference in the reproduction number at the initial phase. (a) The reproduction numbers estimated with the proposed state-space method for 10 days until the day before the lockdown measures of each country. Days are counted from 12 days before the confinement measures. (b) Initial variation in the numbers of daily new cases; *n_i_* divided by *n*_6_. The period is shifted by 5 days, by taking account of the typical transmission delay.

### Future prediction

Using the proposed method, it is also possible to predict the number of new cases in the future. This can be done by simulating the converted Hawkes process Eq. (2) with the parameters estimated from the given data. One may adopt the reproduction number *R_i_* in near future as constant at the value of an endpoint of estimation if the current conditions are assumed to be maintained. Alternatively, one may also examine various time schedules of *R_i_*, by assuming possible choices of relaxation or confinement measures.

In Fig. 6 we applied the forecasting method to the data of Japanese daily cases. Assuming that we are on June 30, 2020, we have estimated the reproduction number 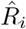 using the daily cases until that day. To predict the number of daily cases from July 1to August 1, we ran the Hawkes process 100 times to obtain the expected daily cases. Firstly, we have assumed that the reproduction number remains the value obtained for the last day *R* = 1.4. Occasionally the reproduction number has not changed drastically in July, and accordingly, the predicted number of new cases is similar to the real data obtained in July.

**FIG. 6.**
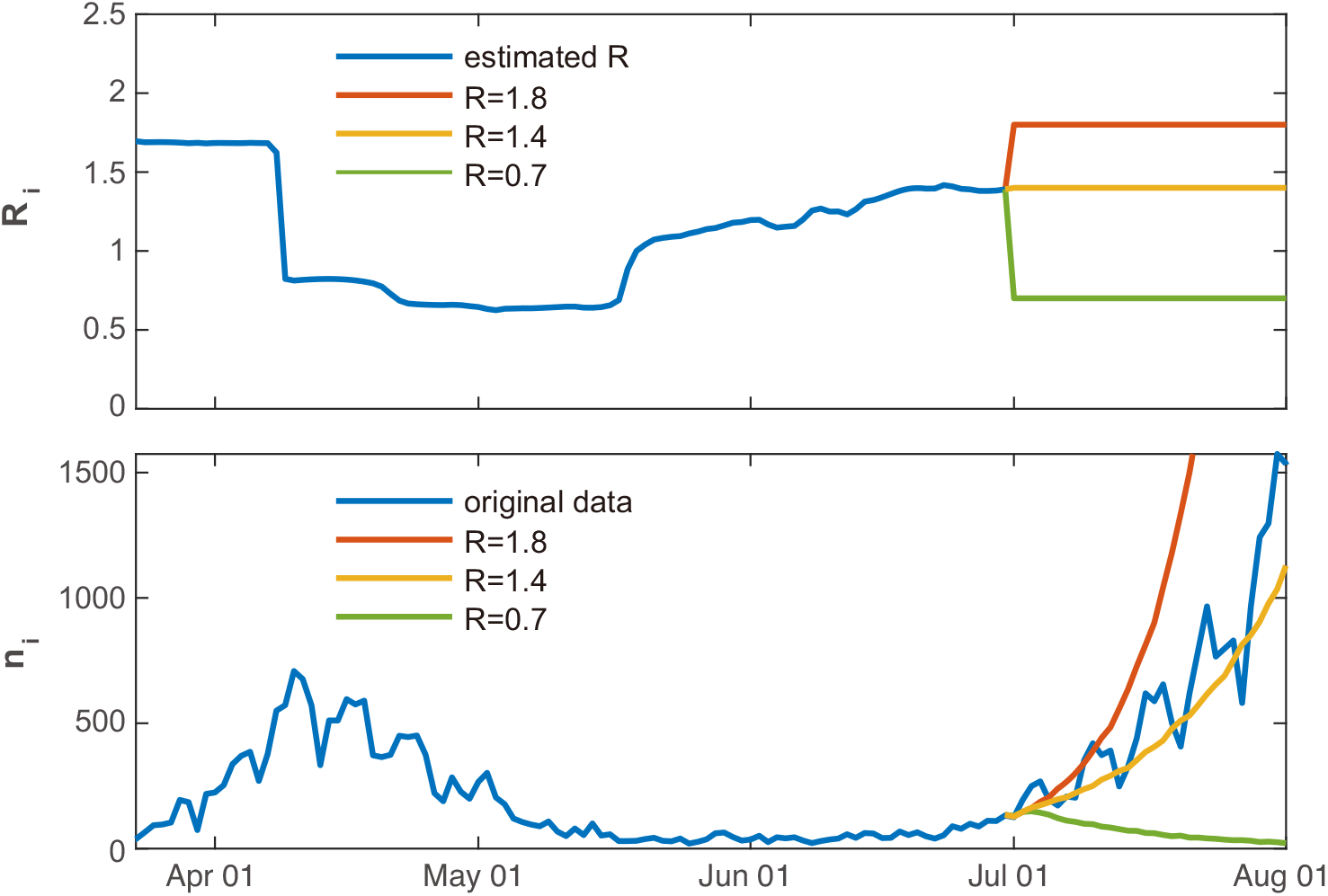
Predicting the number of new cases in the future. The forecasting method was applied to the data of Japanese daily cases, assuming that we were on June 30, 2020. We ran the Hawkes process for 100 times to obtain the expected daily cases, by assuming that the reproduction number remains constant *R* = 1.4, which was obtained using the previous data (orange line). We also examined the cases in which the reproduction number is decreased to *R* = 0.7 due to confinement measures (green line) or increased to *R* = 1.8 by liberalization (red line).

We have also tested the cases in which the reproduction number is decreased to *R* = 0.7 due to confinement measures, or increased to *R* = 1.8 by liberalization. In this way, we may examine what might occur if political interventions are taken.

## DISCUSSION

Society and the media currently alternate between hope and despair in response to the temporary decrease or increase of daily new COVID-19 infections, which came out after a long latency period. To make an objective assessment of the current status, we have developed a state-space method for estimating the control status, in particular quantifying the time-dependent reproduction number *R*.

### Pros and cons

We adopted the Hawkes process, or the self-exciting model, in describing the variables underlying the transmission of disease. In contrast to ordinary differential equation models such as SIR or SEIR models, the Hawkes process is advantageous in that it explicitly specifies the distribution of transmission delays. However, the Hawkes process does not account for the finite size effect, in which infected and recovered people represent a finite fraction among the entire population. There have been some models that incorporate the finite population size effect into the Hawkes process, as has been done with the SIR or SEIR models [18, 19]. To analyze the current status of COVID-19, however, we do not take the finite size effect into account, as the fraction of the recovered or removed people is still less than a few % of the entire population.

While preparing this manuscript, the authors discovered another interesting approach that applied the Hawkes process for modeling COVID-19 transmission [20]. In that study, the authors combined the Hawkes process with spatial and temporal covariates, such as demographic features and Google mobility indices, to explain the variability of the reproduction number, and to forecast future cases and deaths in the USA. By contrast, our state-space method estimates the change-points in the reproduction number solely from a series of daily cases. Though these methods apply the Hawkes process differently, both methods are seen to demonstrate the effectiveness of this model in analyzing epidemics.

We introduced the Cauchy distribution, Eq. (7), into our analysis, assuming the stepwise changes in the reproduction number *R_i_*. Accordingly, we were able to infer change-points from the posterior distribution taking on stepwise characteristics. It should be noted that, however, there may be an additional latency between the times at which political measures were conducted and the changes in the reproduction number, which may reflect the change in behavior. This delay may also be country-specific. Therefore, it could be interesting to investigate the delay in the change-points in the reproduction number following social events.

When inferring the transmission of disease from daily confirmed cases, we have considered potentially erroneous observations made in the real data. We took into account counting errors by assuming a negative binomial distribution that represents the over-dispersion. We also took into account the variation by day of the week and adjusted the data by compensating for the periodic dependency. Note that there may still be an underestimation of infection numbers, as asymptomatic cases may have been overlooked. Though this is unavoidable unless the inspection is enforced, it is reported that the infections caused by asymptomatic people are relatively small (about 6%) for COVID-19 [15].

We have assumed that the transmission delay is a serial interval defined as the duration between symptom onsets of successive cases and adopted the log-normal distribution with the mean 4.7 days and SD 2.9 days, as suggested in reference [11]. As our mathematical formulation is general, it is possible to search for a more suitable transmission kernel *ϕ_d_* without relying on such external knowledge, if the numbers of daily cases are accurately provided.

The most crucial assumption in the majority of mathematical model studies, including this study, is the mean-field assumption, in which all individuals are assumed to interact uniformly. Though difficult to incorporate, it is desirable to consider the heterogeneity of the real-world community in analyzing the communicability of disease.

Despite these assumptions, the proposed state-space method may be of worth in assessing the status of the disease systematically, based on reported daily confirmed cases. This method might serve as a reference for governments adopting variable regulations that should be changed according to current infection circumstances.

## METHODS

We have constructed a state-space method for estimating the time-varying reproduction number by assuming that new cases have been generated with the converted Hawkes process, Eq. (2).

### Deriving the number of events

The number of events *v_i_* or *v* is derived from a distribution specified with the mean rate *λ_i_* or *λ*. It would be natural to assume the Poisson distribution 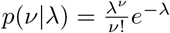. However, real data are subject to erroneous observation and accordingly they tend to be over-dispersed, or the sample variance exceeds the sample mean. Here, we incorporate over-dispersed data using the negative binomial distribution [21]:

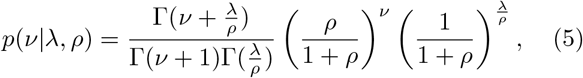

where *ρ (>* 0) represents the degree of over-dispersion, or the variance is Var(*v*) = (1 + *ρ*)*λ*. The Poisson distribution is in the limit of *ρ* → 0.

The degree of over-dispersion of the real data *ρ* can be estimated as described later. The estimated *ρ* turned out to range from 20 to 200 for real data, and accordingly, we have assumed *ρ* = 50 in making the numerical simulations depicted in Fig. 2.

### The system equation

To detect change-points in the reproduction number {*R*_i_}*i* in Eq. (2), we introduce a method of estimating stepwise dynamics [22]. We assume that system’s state *x_i_* obeys the evolution

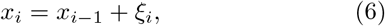

with the Cauchy random number *ξ*:

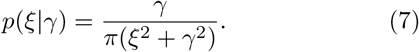

We assume that the reproduction number is given as *R_i_* = *f(x_i_)* with the non-negative function. Here we adopted a ramp function *f*(*x*) = max(0, *x*).

### State inference

We construct a state-space model for estimating the temporally changing reproduction number *R_i_* from a given dataset of daily confirmed cases {*v_i_,…,v_T_*}, which may have been converted from the original data {*n*_1_*,… ,n_T_*} according to Eq. (4). The basic procedure of constructing the state-space method is similar to the one we developed for estimating exogenous and endogenous factors in a chain of point events [23].

To put the model in the state-space form, we take the summation in Eq. (2) over the last *L* days,

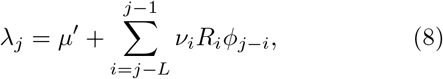

and introduce a concatenated state vector,

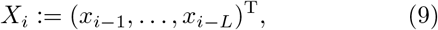

so that the rate process (8) depends only on the current state *X_i_*. Accordingly, the state *X_i_* obeys the evolution

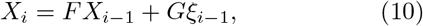

where

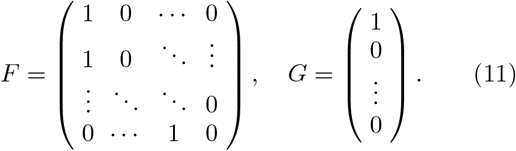

We have chosen *L* = 30 in the following analysis.

The posterior distribution of system’s state *X_i_*, given a set of daily new cases until *i*th day *Y_i_*:= {*v_i_,…, v_i_*} is obtained using Bayes’ theorem as

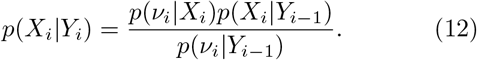

Here, *p*(*X_i_*|*Y_i_*_−1_) may be obtained using a system model *p*(*X_i_*|*X_i_*_−1_) and the posterior distribution on day *i −* 1, *p*(*X_i_*_−1_|*Y_i_*_−1_), as

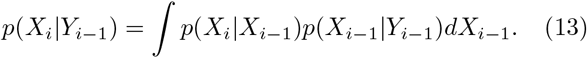

Starting from the initial distribution *p*(*X_i_*|*Y_0_*), we iterate Eqs. (12) and (13) to compute *p*(*X_i_*|*Y_i_*_−1_) and *p(X_i_|Y_i_)* for *i* = 1, 2,…,*T*.

Then, we compute the distribution of system’s states {*X*_1_,*… ,X_T_*}, given an entire set of occurrences *Y_T_*:= {*v*_1_*,…, v_T_*} with

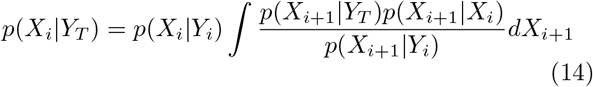

in reverse order as *i* = *T −* 1*, T −* 2*, …*, 1, using the distribution functions *p(X_i_|Y*_i_) and *p(X_i_|Y_i−_*_1_), which were obtained with Eqs. (12) and (13).

We then take the median of the posterior distribution *p*(*X_i_*_+1_|*Y_T_*) for the estimate of the state 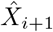. The estimate of the reproduction number, 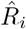, is then given by the first element of 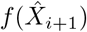. With the estimated reproduction number, we obtain the estimated total rate as

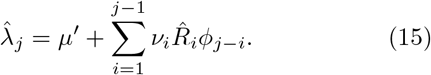

We devised an algorithm that performs the integrations in Eqs. (12), (13), and (14) numerically using a sequential Monte Carlo method [24, 25].

To avoid bias in estimating the state, which is caused by outliers in the data, we may discard the preassigned outliers and treat them as “missing observations” [25], for which the posterior distribution of *X_i_*, conditional on *Y_i_*, is set to *p(X_i_|Y_i_)* = *p(X_i_|Y_i−i_*) without applying the Bayesian update (12).

The state-space method possesses a hyperparameter *γ* that characterizes the Cauchy distribution of the system equation Eq. (7). The hyperparameter was determined by applying the state-space method to synthetic data shown in Fig. 2. We tested different values for the hyperparameter *γ* as 10^−2^, 10^−3^, and 10^−4^, and confirmed that the free energy was lowest for the case of *γ* = 10^−3^. Accordingly, we fixed the hyperparameter at *γ* = 10^−3^.

The over-dispersion parameter *ρ* can be determined as

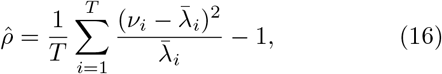

where 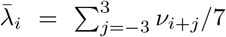 represents the mean daily cases averaged over a week.

With the hyperparameter *γ* = 10^−3^ and the parameter 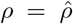 determined for each dataset with Eq. (16), we perform the sequential Monte Carlo algorithm with 10^6^ particles to compute the posterior distributions of the reproduction number for each day.

### Code availability

The application program and example datasets are available at our website https://s-shinomoto.com/COVID/ and the site hosted publicly on GitHub, accessible via https://github.com/shigerushinomoto.

## Data Availability

https://github.com/shigerushinomoto

## APPENDIX: APPLYING THE CONVENTIONAL WT METHOD TO REAL DATA

Figure 7 compares the proposed method with a conventional WT method [17] by applying them to the real data examined in Fig. 4. Similar to the analysis of synthetic data in Fig. 2, the WT method is easily influenced by the fluctuation of data. Though both methods indicate similar values, our method is robust to the large fluctuations in the data and can indicate possible change points.

**FIG. 7.**
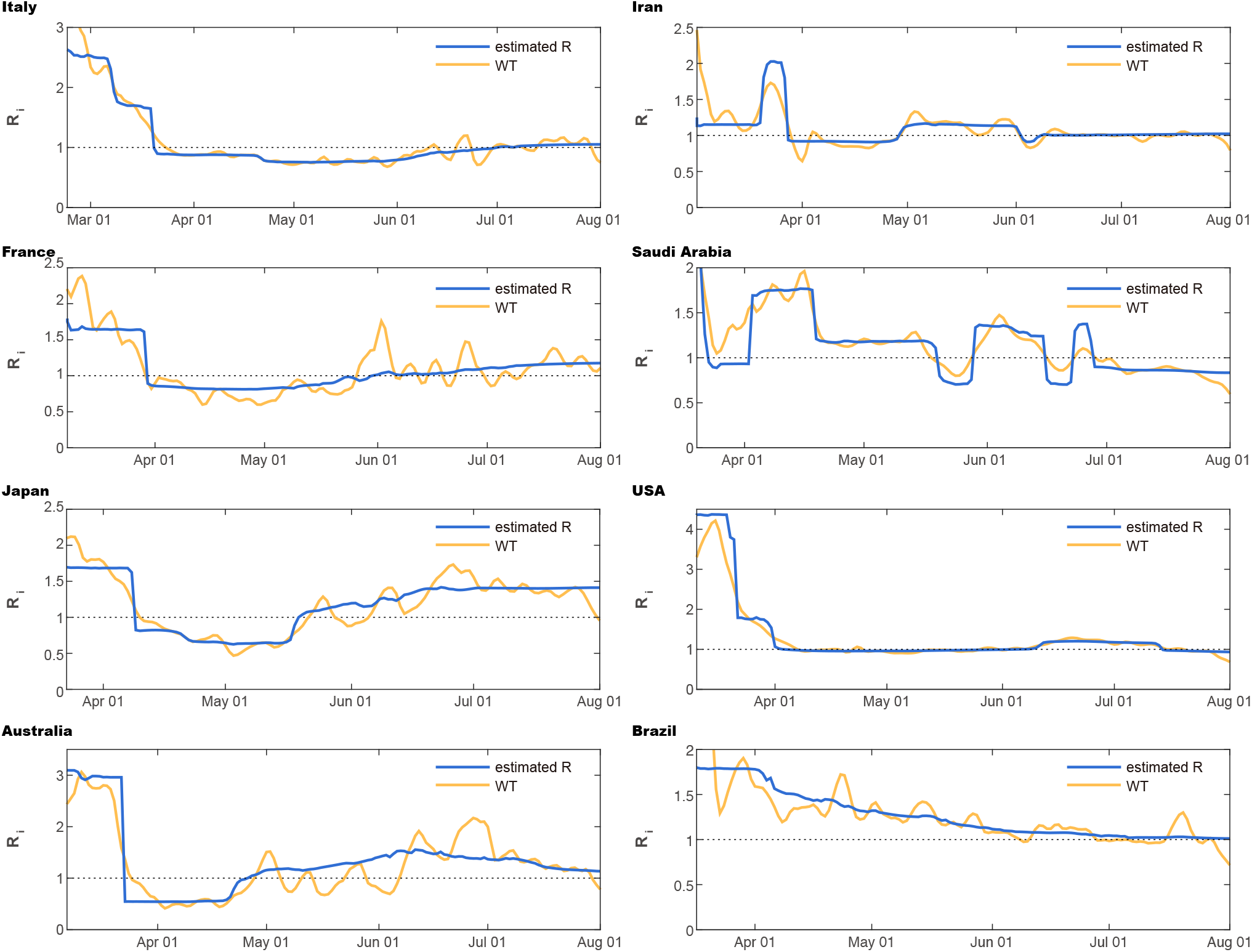
Comparison of the state-space method (blue line) and the conventional WT method (orange line), when applied to real data in Fig. 4. (left) Italy, France, Japan, and Australia; (right) Iran, Saudi Arabia, and the USA, and Brazil.

## ACKNOWLEDGMENT

We thank Shin Takagi, Masaki Ogura, Ryota Kobayashi, Takaaki Aoki, and Hideaki Shimazaki for their constructive comments on this manuscript, and Hidetaka Manabe for his technical assistance in developing a web-application program. S.S. is supported by the New Energy and Industrial Technology Development Organization (NEDO).

## Notes

### Competing Interest Statement

The authors have declared no competing interest.

### Author Declarations

This is not applicable to our study, because our paper represents a method of analyzing the number of COVID-19 infections that is publicly available.

### Summary of Updates

Figures 2 and 4 revised; New analysis and Figures 6 and 7 added.

